# Human Papillomavirus Infection Rate by Genotype and Vaccination Rates in Canada: Canadian Health Measures Survey 2009 to 2013

**DOI:** 10.1101/2022.04.05.22273469

**Authors:** Yi-Sheng Chao

## Abstract

**Background:** An infection with certain HPV genotypes can lead to cancer or genital warts. HPV can be detected with PCR-based tests, and some genotypes can be prevented by vaccines. However, since the infection rates of various HPV genotypes have not been well reported, the present study aims to provide this information.

**Methods:** The Canadian Health Measures Survey (CHMS) is an ongoing biannual national survey. Between 2009 and 2011, it sampled a nationally representative sample of females aged 14 to 59 years to determine the infection rates of 46 HPV genotypes. Females aged 9 to 29 years and 9 to 59 years were asked whether they received HPV vaccines between 2009 to 2011 (cycle 2) and 2012 to 2013 (cycle 3), respectively. The reported infection rates and vaccination proportions were weighted and adjusted for the survey design.

**Results:** Among the estimated 10,592,968 females aged 14 to 59 years at cycle 2, the HPV genotypes with the highest infection rates were 16, 62, 74, and 54, and the rates were 3.42% (95% CI = 1.67% to 5.17%), 2.14% (95% CI = 0.68% to 3.59%), 2.1% (95% CI = 0.51% to 3.69%), and 2.04% (95% CI = 0.38% to 3.7%), respectively. There were an estimated 6,569,100 and 11,603,752 females aged 9 to 29 and 9 to 59 years at cycles 2 and 3, respectively. The proportions receiving a HPV vaccine were 13.55% (11.18% to 15.92%) and 12.3% (9.8% to 14.79%), respectively. The estimated numbers of females that received HPV vaccines were 890,197 and 1,427,000, respectively.

**Conclusion:** Canada is one of the few countries that conduct national surveys to determine HPV infection rates by genotype, which are not limited to the surveillance of carcinogenic genotypes. Our study found discrepancies between the HPV genotypes whose infections were the most common, that could be detected by PCR tests, that were carcinogenic, and that could be prevented by vaccines. For example, 5 of the 7 genotypes (42, 54, 62, 66, and 74) with infection rates of more than 1% cannot be detected by PCR tests and are not targeted by vaccines. HPV 51 is carcinogenic, associated with genital warts, and can be detected by PCR tests, but it is not targeted by vaccines. We recommend a better alignment of the genotypes targeted by HPV tests and vaccines with those genotypes with the highest infection rates in Canada.

## Introduction

Human papillomavirus (HPV) infection is a major cause of cervical cancer and is related to other cancer or non-cancer lesions.[1, 2] The risk of cancer depends on the HPV genotypes.[1, 2] Figure 1, based on the findings of the International Agency for Research on Cancer (IARC), identifies 12 HPV genotypes as carcinogenic (HPVs 16, 18, 31, 33, 45, 52, 58, 35, 39, 51, 56, and 59) and 1 other as probably carcinogenic (HPV 68).[1, 2] Seven of the 13 genotypes (HPVs 16, 18, 31, 33, 45, 52, and 58) are associated with the occurrence of more than 90% of invasive cervical cancer.[3] HPV 6 and 11 are associated with more than 90% of genital warts.[4] The prevalence of HPV infection differs by countries. Globally, in 2012, HPV infection was reported to be prevalent among 10% of healthy women.[5] The infection rate ranges from 6.6% in Europe to 22.9% in Africa, and HPV 16 infection is the most common among all HPV genotypes.[5] In Canada, HPV is estimated to be the most common sexually transmitted disease, and more than 70% of sexually active Canadians are expected to have HPV infection in their lifetime.[6] It is estimated that 6.2% of individuals with normal cervical cytology were infected with HPV 16 or 18.[7, 8] In terms of infection detection, polymerase chain reaction (PCR) based tests can detect at least 13 high-risk genotypes (16, 18, 31, 33, 45, 52, 58, 35, 39, 51, 56, 59, and 68) and are used for cervical cancer screening.[8] Those individuals identified with active HPV infection may be referred to cervical exams for confirmation.[8] Currently, HPV tests have not been included in routine cancer screening programs in Canada.[1]

**Figure 1.**
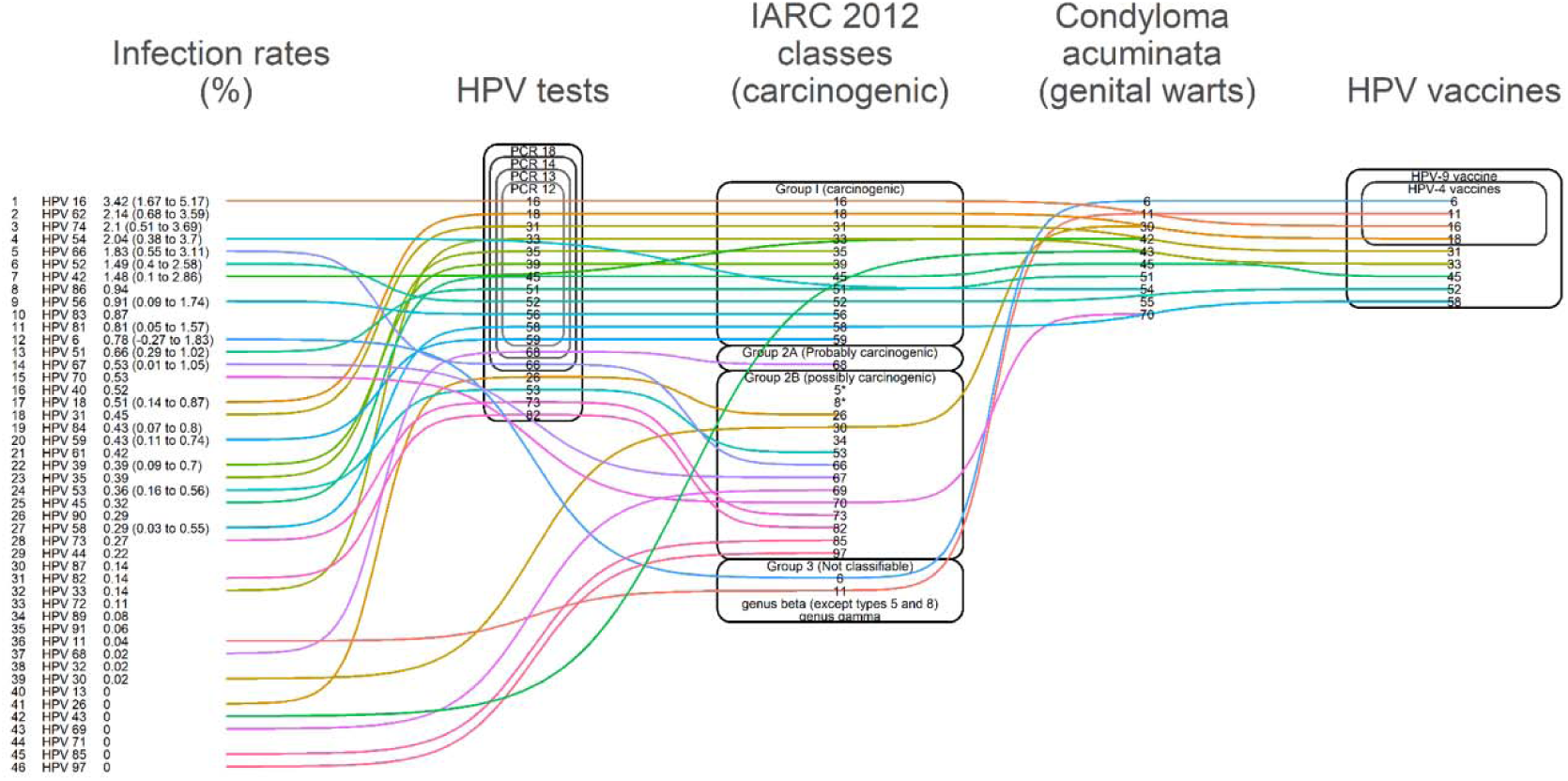
HPV genotypes by infection rates, HPV tests, carcinogenicity, association with genital warts, and HPV vaccines. *In patients with epidermodysplasia verruciformis. HPV = human papillomavirus; IARC = International Agency for Research on Cancer; PCR = Polymerase chain reaction. Infection rates are obtained from a national representative sample of females aged 14 to 59 years participating the Canadian Health Measures Survey. The HPV genotypes are listed in descending order of infection rates. The HPV tests include the 13, 14, 15, and 19 HPV genotypes that are targeted by diagnostic tests using PCR.[1] The IARC classes are obtained from the IARC database (https://monographs.iarc.fr/list-of-classifications).[8] The HPV genotypes associated with Condyloma acuminata (genital warts) were obtained from a review.[38] The HPV vaccines include the 4 and 9 HPV genotypes that these vaccines aim to prevent.[10] The same HPV genotypes are connected to each other across the categories.

Vaccination is an important strategy to prevent HPV infection[7, 9], and HPV vaccines have been approved in Canada since 2006.[10] Between 2007 and 2010, public vaccination programs began to be implemented for girls in various Canadian jurisdictions.[10] These public vaccination programs have been expanded and now are available to other populations.[10] Several HPV genotypes are targeted by vaccines, especially 6, 11, 16, and 18.[10] Also, other vaccines can be used to additionally target HPV 31, 33, 45, 52, and 58.[10] In Canada, the vaccination rates found by various studies are estimated to be 69.62% and 18.66% for in-school programs and community-based programs, respectively.[4] The vaccination rates for girls eligible for HPV vaccination programs vary to a great extent between Canadian jurisdictions, from 40% in the Northwest Territories to 91% in Newfoundland.[11]

HPV genotypes differ with respect to the development of various lesions, as to whether they can be detected by available HPV tests, and as to whether they can be prevented by vaccination. The infection rates of different HPV genotypes provide important information for comparison with the assessment of potential disease burden, detectability via PCR tests, and vaccine effectiveness. However, the estimates for HPV infection rates and vaccination rates are based on sporadic reports and diverse data sources. Such information is crucial for assessing the effectiveness of public vaccination programs. Based on nationally representative Canadian data, the present study reports on HPV infection rates by genotype and HPV vaccination rates.

## Methods

The Canadian Health Measures Survey (CHMS) is an ongoing biannual national biomonitoring survey that has interviewed Canadians since 2007.[12-16] Canadians who were full-time military members, reserve residents, and institutional residents—less than 4% of the population—were excluded from the sampling.[16, 17] More than 5,000 participants were interviewed in each of the 5 cycles that released data before Dec 2020.[12, 15-17] In addition to face-to-face interviews, biological samples were obtained from the participants to determine levels of various biomarkers or chemicals.[12-15, 18] By adjusting for the sampling strategies and bootstrap weights, about 30 million Canadians were estimated for each of the 5 cycles, and about half of them were female.[12, 14, 15, 17] Population characteristics and the numbers of population by age were published elsewhere.[12, 14, 15, 19, 20]

In cycles 2 and 3, conducted between 2009 to 2011 and 2012 to 2013, respectively, participants were asked whether they had received a HPV vaccine.[21] In cycle 2, 1,576 female respondents aged 9 to 29 years were asked[21], and in cycle 3, 1,845 female respondents aged 9 to 59 years were asked.[21] The vaccination rates among eligible Canadians were reported in the survey.

In cycle 2, 46 HPV genotypes were tested using urine samples (6, 11, 13, 16, 18, 26, 30, 31, 32, 33, 35, 39, 40, 42, 43, 44, 45, 51, 52, 53, 54, 56, 58, 59, 61, 62, 66, 67, 68, 69, 70, 71, 72, 73, 74, 81, 82, 83, 84, 85, 86, 87, 89, 90, 91, and 97) (see Figure 1).[22] Samples were obtained from 1,770 female respondents aged 14 to 59 years.[21] The infection rates among eligible Canadians were reported in the survey.

## Data analysis

To adjust for the survey design, bootstrap weights, sampling strata, regions, and personal weights were used in the data analysis.[12-15, 23, 24] To protect the survey respondents’ confidentiality, the results were vetted based on specific rules, before they were released by the Research Data Centre to researchers.[25] Only weighted statistics were released[25], and they needed to meet a minimum sample size requirement.[26-30] To protect participant confidentiality and prevent participants from being identified using the released data, Statistics Canada did not allow the release of calculated proportions based on 10 or fewer unweighted participants. At least one participant in each sampling site for each cycle was required.[12-15] In total, to meet the minimum sample size requirement, roughly at least 100 respondents were required for each cycle.[12-15] For these proportions that did not meet the requirement, 95% CIs could not be released, and so the proportions provided in CHMS data dictionaries were used.[22] All data analysis was conducted with R (v3.20)[51] and RStudio (v0.98.113).[52]

This secondary data analysis was approved by the ethics review committee at the Centre Hospitalier de l’Université de Montréal. A consent to participate was not required for this secondary data analysis.

## Results

Cycle 2 (2009 to 2011) estimated 10,592,968 female Canadians aged 14 to 59 years. The HPV genotypes with the highest infection rates were 16, 62, 74, and 54, and the rates were 3.42% (95% CI = 1.67% to 5.17%), 2.14% (95% CI = 0.68% to 3.59%), 2.1% (95% CI = 0.51% to 3.69%), and 2.04% (95% CI = 0.38% to 3.7%), respectively. Three HPV genotypes (66, 52, and 42) were estimated to have infection rates between 1% and 2%. The other 39 HPV genotypes were estimated to have infection rates less than 1%. The unweighted numbers of participants infected with 28 HPV genotypes were fewer than those required by Statistics Canada, and so 95% CIs of the infection rates of these HPV genotypes could not be released. Thus, the infection rates of these genotypes were derived from data dictionaries (11, 13, 26, 30, 31, 32, 33, 35, 40, 43, 44, 45, 61, 68, 69, 70, 71, 72, 73, 82, 83, 85, 86, 87, 89, 90, 91, and 97). As a result, 95% CIs of the infection rates were not determined. No infection was identified with 7 HPV genotypes (13, 26, 43, 69, 71, 85, and 97).

### Vaccination rates

Cycle 2 (2009 to 2011) and cycle 3 (2012 to 2013) estimated 6,569,100 and 11,603,752 female Canadians aged 9 to 29 and 9 to 59 years, respectively. The proportions receiving an HPV vaccine were 13.55% (95% CI = 11.18% to 15.92%) and 12.3% (95% CI = 9.8% to 14.79%), respectively. The numbers of females receiving HPV vaccines with respect to the respective age ranges at cycles 2 and 3 were 890,197 and 1,427,000, respectively.

### Infection rates, detection, carcinogenicity, and preventability

In Figure 1, HPV 16 is carcinogenic,[8] can be detected by PCR-based HPV tests,[31] and can be prevented by HPV vaccines.[10] However, HPV 16 is not related to genital warts and is not linked to the genotypes associated with genital warts. In contrast, 5 of the 7 HPV genotypes (42, 54, 62, 66, and 74) with infection rates of more than 1% cannot be detected by PCR-based tests (13 genotypes)[31] and are not targeted by HPV vaccines.[10]

There were 13 genotypes with infection rates less than 1% (HPVs 32, 35, 40, 44, 61, 72, 81, 83, 84, 87, 89, 90, and 91) that are not carcinogenic, cannot be detected by PCR tests, are not associated with genital warts, and are not targeted by vaccines. HPVs 6 and 11 are associated with genital warts and are targeted by vaccines, but they are not carcinogenic and cannot be detected by PCR tests. HPV 51 is carcinogenic, associated with genital warts, and can be detected by PCR tests, but it is not targeted by vaccines. HPVs 35, 39, 56, and 59 are carcinogenic and can be detected by PCR tests, but they are not associated with genital warts and are not targeted by vaccines. HPVs 53, 68, 73, and 82 can be detected by specific PCR tests (19 genotypes), but they are not carcinogenic, are not associated with genital warts, and are not targeted by vaccines. HPVs 18, 31, 33, 52, and 58 can be detected by PCR tests (13 genotypes), are carcinogenic, and are targeted by vaccines (4, 9, 9, 9, and 9 genotypes, respectively), but they not associated with genital warts. HPV 45 can be detected by PCR tests (13 genotypes), is carcinogenic, and is associated with genital warts, and is targeted by specific vaccines (9 genotypes). HPV 70 is associated with genital warts, but it is not carcinogenic, cannot be detected by PCR tests, and is not targeted by vaccines. HPV 30 is possibly carcinogenic and is associated with genital warts, but it is not carcinogenic and is not targeted by vaccines.

Among the HPV genotypes without any identified infection, HPVs 13 and 71 are not detected by PCR tests, are not carcinogenic, are not associated with genital warts, and are not targeted by vaccines. HPV 26 can be detected by specific PCR tests (19 genotypes), but is not carcinogenic, is not associated with genital warts, and is not targeted by vaccines. HPV 43 is associated with genital warts, but it is not detected by PCR tests, is not carcinogenic, and is not targeted by vaccines. HPVs 69, 85, and 97 are possibly carcinogenic, but they cannot be detected by PCR tests, are not associated with genital warts, and not targeted by vaccines.

## Discussion

The present study reports the HPV infection rates by genotype determined at a time when HPV vaccines were beginning to be used increasingly and were being promoted by public programs in Canada. Such information is crucial for assessing the epidemiology of HPV infection by genotype and the effectiveness of HPV vaccines. More importantly, we identified discrepancies between the HPV genotypes whose infections are prevalent, that can be detected by HPV tests, that are associated with cancer or genital warts, and that are targeted by vaccines. Since the HPV genotypes considered carcinogenic may change over time, depending on available evidence,[32-35] the usefulness of commercially available PCR tests approved in Canada—especially those reviewed by Health Technology Assessment (HTA)—may be somewhat limited with respect to the genotypes chosen for HPV vaccines, and the choice of targets for HPV infection surveillance.

### Genotypes with high infection rates

HPV 16 is carcinogenic[8] and is the most common HPV infection in the world.[5] It also is the genotype with the highest infection rates in Canada, a fact that has been recognized by other researchers.[5, 36] The estimated infection rates of HPV 16 varies from 3.5% in healthy women in North America[5] to 4.7% of the normal cytology[36] in Canada. The present study estimates that between 2009 and 2011, 3.42% of Canadian females aged 14 to 59 years were infected with HPV 16. However, studies disagree about the other HPV genotypes that have high infection rates and the percentage of their infection rates. One study has estimated that HPVs 53 and 52 are the genotypes with the second and third highest infection rates in healthy women in North America.[5] In this study, four out of the five HPV genotypes studied were carcinogenic (HPVs 16, 18, 39, and 52), and the other one (HPV 53) was possibly carcinogenic.[5] In another study that systematically reviewed and meta-analyzed trials or studies in Canada, HPVs 18 and 31 were the genotypes with the second and third highest infection rates.[36] In this study, 9 out of the 10 HPV genotypes studied were carcinogenic, and the other one (HPV 66) was possibly carcinogenic.[36] Results from previous studies may suggest that researchers’ interests were focused mostly on carcinogenic HPV genotypes, so infections from other HPV genotypes may not have been as well studied as they should have been. The present study found that the infections of several possibly carcinogenic genotypes (especially HPV 66) were more prevalent than many carcinogenic HPV genotypes.

The infection rates of carcinogenic HPV genotypes were mostly lower than those rates estimated by synthesizing primary studies of various characteristics. For example, based on a 2014 meta-analysis, HPVs 18, 31, 66, 45, and 59 (ordered by infection rates) were the carcinogenic genotypes estimated to be prevalent in more than 1% of normal cytology.[36] However, in the nationally representative sample of females aged 14 to 59 years surveyed in the CHMS, the infection rates of these genotypes were 0.51%, 0.45%, 1.83%, 0.32%, and 0.43%, respectively. It is unclear whether the included smaller studies tended to obtain more high-risk individuals or whether small-study effects have been well addressed.[37] The reasons for the discrepancies in the estimated infection rates between studies were not clear. Thus, an extensive review may be required of the population characteristics between studies and follow-ups of infection trends in future cycles of the CHMS.

A lack of detection methods or preventive measures for certain HPV genotypes with the highest infection rates does not mean these genotypes are not associated with adverse health outcomes. Among the genotypes that could not be detected by PCR tests (12 to 18 genotypes) in a health technology assessment (HTA) review, HPVs 42, 62, and 74 were associated with the development of cervical lesions, cervical intraepithelial neoplasia.[38] The PCR tests reviewed by the HTA were the tests that are used in primary screening, and many have been approved for clinical use in Canada.[31] In Figure 1, HPVs 42 and 54 are associated with genital warts.[38] We did not identify the major studies that have investigated the health impact of these genotypes with high infection rates in Canada.

### Detection and carcinogenicity

The HPV genotypes that PCR tests aim to detect include the 12 carcinogenic genotypes identified by the IARC classification updated in 2012.[31] HPVs 26, 53, 66, 68, 73, and 82 are the genotypes that can be detected using the PCR tests that can detect up to 18 genotypes.[31] In the IARC 2012 update, HPV 68 is considered probably carcinogenic, and HPVs 26, 53, 66, 73, and 82 are possibly carcinogenic.[33] HPVs 26 and 73 also are associated with common skin lesions.[38]

However, nine possibly carcinogenic genotypes (HPVs 5, 8, 30, 34, 67, 69, 70, 85, and 97) cannot be detected with the PCR tests (12 to 18 genotypes) reviewed by the HTA. In Figure 1, the HPV genotypes associated with genital warts cannot be detected by the PCR tests that can identify 12 to 18 genotypes.[31] Moreover, vaccine-preventable HPV genotypes (6 and 11) cannot be detected by these PCR tests. The role of these commercially available PCR tests may be limited in detecting some possibly carcinogenic genotypes and those associated with genital warts. With respect to policy evaluation, the genotypes that are not targeted by these PCR tests should be considered when assessing HPV-related disease burden or the effectiveness of preventive measures.

Moreover, one major limitation of many commercially available PCR tests is that only positive or negative results are reported, and the exact HPV genotypes detected are not reported.[31] If the HPV genotypes can be reported by PCR tests, this may help efforts to prevent further spreading of these genotypes.

### Changes to the carcinogenicity classification

The 12 genotypes that can be detected by PCR tests match those classified carcinogenic genotypes identified in the IARC 2012 update.[33] Figure 1 shows the additional genotypes that can be identified by PCR tests that can detect 13, 14, or 18 genotypes that are probably or possibly carcinogenic as defined by the IARC. Over time, changes have been made to the carcinogenicity classifications of HPV genotypes.[35, 39] Although the present study found that the infection rates of the possibly carcinogenic genotypes were not high, except for HPV 66, their potential for carcinogenicity requires constant surveillance. Since some genotypes have high infection rates, it may be easier to identify the patients infected with these genotypes for follow-up. The CHMS can be used to assess related changes in the cancer burden attributable to HPV infection, and which is preventable by vaccines.[40] The present study did not consider several possibly carcinogenic genotypes (HPVs 5, 8, and 34) that may need follow-up in future CHMS cycles.

### Vaccination rates

The vaccination rates reported in the two CHMS cycles were based on two different age ranges—9 to 29 and 9 to 59 years—and these rates decreased slightly over the two cycles from 13.55% to 12.3%, respectively. However, the absolute numbers of females receiving the vaccines over the two CHMS cycles increased more than 60%. Vaccination programs in Canada are expanding with public support.[9, 11] The target population also changed from school-age girls in many provinces to other age groups and males.[41] Globally, between 2006 and 2019, HPV vaccination indicated an increasing trend, and Canada was one of the countries with more than 83% coverage among school-aged girls.[42] However, a considerable variability exists in HPV vaccination rates across Canada.[42] In a recent estimate, the coverage rates of school-based HPV vaccination programs ranged from 59.4% in Ontario to 88.4% in Prince Edward Island for girls in the 2016–2017 school year, compared to 52.5% in Ontario to 88.2% in Newfoundland and Labrador for girls in the 2008–2009 school year.[43] Continuous efforts to vaccinate younger generations will lead to better vaccination coverage.

It has been estimated that HPV vaccines can reduce cervical cancer and other HPV-related cancers.[40] However, since most vaccines act against four or nine HPV genotypes,[10] many of the genotypes with high infection rates in this study were not targeted. Nevertheless, some evidence exists that cross-reactivity may be induced by HPV vaccines to act against non-genital HPVs.[44] At a population level, the herd immunity that protects unvaccinated individuals also can be achieved when more individuals are vaccinated.[45]However, the exact impact on infection rates of the genotypes not targeted by vaccines remains unclear. We expect vaccination rates to continue increasing, and the influence of vaccines on the infection rates of genotypes not targeted needs to be monitored.

### Future research directions

The HPV genotypes with the highest infection rates provide opportunities for researchers to assess their health impact using human observational data. Carcinogenicity classification depends not only on animal data, but also on human trials or studies.[32] By following up individuals infected with HPV, it is possible to understand the impact of these HPV genotypes on health. The present study found that HPVs 54, 62, 74, and 86 are common and may be good candidates for long-term health studies.

Although these HPV genotypes are not directly targeted by vaccines, studies have shown that cross-reactivity with non-genital HPVs may be induced by vaccines.[44, 46] However, it is unclear whether cross-reactivity may be induced against the HPV genotypes with high infection rates. In the future, the impact of HPV vaccines on genotypes they target or do not target may be evaluated by using the data generated by large studies like the CHMS.

Over time, the use of HPV vaccines has been expanded to other age groups and males.[10] Thus, in the future, it is important to collect data not only on other HPV genotypes, but also on other population groups. The vaccinations against four or nine genotypes may cause spill-over effects or herd immunity on other genotypes[44] or population groups. The infection rates and vaccination history of other population groups may interact with the effectiveness of cervical cancer prevention. Long-term follow-up may be needed. With the expansion of vaccination programs, the cost effectiveness analysis of using HPV tests for primary cervical cancer screening[31] may need to be updated.

IARC HPV Prevalence Surveys conducted outside of Canada have reported a clustering of particular HPV genotypes.[47] With this knowledge in mind, we are planning to study whether HPV clustering patterns may differ in Canada.

A study has found that the HPV PCR tests reviewed by a HTA did not have perfect diagnostic test accuracy, i.e., 100% sensitivity and 100% specificity, for the detection of cervical cancerous lesions or cervical intraepithelial neoplasia (CIN).[2] Most of the PCR tests reviewed can detect 12 carcinogenic genotypes and 1 probably carcinogenic genotype, and report positivity if any one of these genotypes is detected.[31] Figure 1[8] shows that several possible carcinogenic genotypes and many genotypes associated with CIN[38] are not detected by currently available commercial PCR tests. It is unclear to what degree the lack of detection of possibly carcinogenic genotypes and those genotypes associated with CIN contributes to imperfect diagnostic test accuracy.

Last, herd immunity disturbs HPV infection in a population, and thus protects those who are unvaccinated. In the future, the benefits of HPV vaccination are likely to be seen in the reduction of the incidence of both genital and non-genital cancer.[45] The assessment of the magnitude of these benefits will depend on further data collection and analysis.

### Limitations

The present study has the strength of a single data source, the CHMS, which reported vaccination status and which used consistent methods for HPV genotype quantification. Other studies may have depended on various sources to estimate infection rates, and so it is likely that many HPV genotypes were not tested and reported in the primary studies they reviewed.[36] However, the age ranges for vaccination status and HPV genotyping differed in the CHMS, which leads to some difficulties in assessing the relationship between vaccination status and HPV infection by genotype.

Regarding a subgroup analysis, it is unlikely that Statistics Canada will grant its permission to release the infection rates or vaccination rates of subgroups that did not include large sample sizes. For example, it is unlikely that sufficient sample sizes exist for age-specific infection rates or city-wide infection rates. Moreover, the rates reported may not be applicable to subpopulations of specific characteristics. Only national rates are reported in this study.

The CHMS did not test for more than 46 HPV genotypes and several other genotypes associated with human diseases. For example, it did not test for HPVs 1, 2, and 4 that can cause common skin warts.[5, 38] It seems that the CHMS tested the HPV genotypes that were alpha types only.[5] Yet, it is unclear why these 46 genotypes were tested. We did not identify any CHMS documents that explain why these genotypes were selected for testing. Based on our results, we think strategies for choosing genotypes for surveillance can be refined.

## Conclusion

Canada is one of the few countries that conducts national surveys to determine HPV infection rates by genotype, and it does not limit its surveillance to carcinogenic genotypes.[47] The HPV genotypes with highest infection rates among females aged 14 to 59 years are HPVs 16, 62, 74, 54, 66, and 52 (3.42%, 2.14%, 2.1%, 2.04%, 1.83%, and 1.49%, respectively). With the age ranges expanded from 9 to 29 years to 9 to 59 years between 2009 to 2011 and 2012 to 2013, respectively, 890,197 and 1,427,000 females, respectively, were vaccinated against HPV in the two time periods of the two CHMS cycles. Discrepancies exist between the HPV genotypes whose infections are the most common, that can be detected by PCR tests, that are carcinogenic, and that can be prevented by vaccines.

For example, HPV 16 is carcinogenic, can be detected by PCR tests, can be prevented by vaccines, and is the most common infection among all genotypes. However, five of the seven genotypes (42, 54, 62, 66, and 74) with infection rates more than 1% cannot be detected by most commercially available PCR tests[2, 31] and are not targeted by vaccines. HPVs 6 and 11, whose infection rates were less than 1%, are associated with genital warts and can be prevented by vaccines, but cannot be detected by most commercially available PCR tests.[31] HPV 51 is carcinogenic, associated with genital warts, and can be detected by most commercially available PCR tests,[31] but is not targeted by vaccines. Researchers are developing PCR tests to detect more HPV genotypes[31] and are proposing methods to produce vaccines that can prevent HPV of other genotypes.[48] Currently, some genotypes with the highest infection rates in Canada are not targeted by PCR tests or vaccines. We recommend a better alignment of the genotypes targeted by HPV tests and vaccines with those genotypes with the highest infection rates in Canada.

## Data Availability

It is against the Statistics Act of Canada to release the CHMS data. The collection of the CHMS data has been approved by the ethics committee within the governments of Canada. The CHMS data have been de-identified and maintained by Statistics Canada. The data can be accessed through the Research Data Centres administered by Statistics Canada. The details and eligibility for obtaining data access can be found online (https://www.statcan.gc.ca/eng/rdc/process).

## Declaration

### Required statement for the analysis of Statistics Canada data at the Research Data Centre

The analysis presented in this paper was conducted at the Quebec Interuniversity Centre for Social Statistics (QICSS), which is part of the Canadian Research Data Centre Network (CRDCN). The services and activities provided by the QICSS are made possible by the financial or in-kind support of the Social Sciences and Humanities Research Council (SSHRC), the Canadian Institutes of Health Research (CIHR), the Canada Foundation for Innovation (CFI), Statistics Canada, the Fonds de recherche du Québec - Société et culture (FRQSC), the Fonds de recherche du Québec - Santé (FRQS) and the Quebec universities. The views expressed in this paper are those of the authors, and are not necessarily those of the CRDCN or its partners[25].

### Consent for publication

The consent for publication was not required for this secondary data analysis.

## Acknowledgements

Not applicable.

## Authors’ contribution

YSC conceptualized and designed this study, managed and analyzed data, and drafted the manuscript.

